# Effects of COVID-19 on the provision of Water, Sanitation, and Hygiene (WASH) services in health care facilities in the Far-North Region of Cameroon

**DOI:** 10.1101/2024.02.02.24302188

**Authors:** Carole Debora Nounkeu, Giequel Corniche Noumbissi Massop, Donato Koyalta, Chanceline Bilounga Ndongo, Florent Kamkumo Ouabo, Bertin Nono, Marie Nicole Ngoufack, Jigna Morarji Dharod, Catherine Juillard, Alain Chichom-Mefire, Georges Nguefack-Tsague

## Abstract

Worldwide, the Joint Monitoring Program reports that one in four health care facilities (HCFs) lack functional water supply on premises, one in three lack hand hygiene facilities, and one in three lack adequate infectious waste disposal. The COVID-19 pandemics shed light on the lack of investments, the absence of infrastructures, education and policies related to WASH as well as revealed insufficient investment in healthcare safety and has brought WASH services as non-negotiable for HCFs. This study used a cross-sectional pre-post COVID-19 framework to determine the proportion of HCFs: meeting basic WASH services and, which WASH services improved post-COVID-19 in the Far-North Region of Cameroon. A total of 97 (23.04%) HCFs among the 421 that are found in the Far-North region were surveyed and located in eight (25%) of the 32 Health Districts. They corresponded to the integrated health centers category (79.4%) and the survey’s respondent was the chief of the HCF (92.8%). Approximately 75.3%, 0.0%, 48.5%, 46.4%, and 6.2% of HCFs respectively met thresholds for basic water, sanitation, hygiene, waste management, and environment cleaning services. When comparing pre- vs. post COVID-19 periods, a significant increase (8%) was noted in the proportion of HCFs as of optimal handwashing practices-related services (P=0.0026). There was also a significant increase (p=0.007) in the proportion of HCFs with cleaning protocols available. Further, none of the HCFs fulfilled all the criteria to meet basic services for all the five WASH services. In conclusion, the response to the COVID-19 pandemics only partially improved WASH services-related infrastructures in HCFs of the Far-North Region of Cameroon. The COVID-19 pandemics was a missed opportunity to strengthen WASH services. There should be a continuing encouragement of governments and funding agencies in planning and budgeting WASH in healthcare-related research and issues, and enabling the maintenance of existing WASH infrastructures in healthcare settings.

## Introduction

Health care facilities (HCFs) are recognized and defined by the World Health Organization (WHO) as “environments with a high prevalence of infectious disease agents where patients, staff, caregivers and neighbors of the health-care setting face unacceptable risks of infection if environmental health is inadequate.” They require infrastructures that support water, sanitation and hygiene (WASH) as well as healthcare waste management practices, in order to prevent the spread of disease not only within the HCF, but also to the surrounding community, ensuring quality of care and patients’ safety [1,2]. WASH in HCFs refers to safe and accessible water supply, clean and safe sanitation facilities, hand hygiene facilities at points of care and at toilets, and appropriate waste disposal systems [2,3]. The WHO and the United Nations International Children Emergency Fund (UNICEF) joint monitoring program (JMP) has developed a set of harmonized indicators for WASH in HCFs, corresponding to service levels—basic, limited, and no service—that are used, both to describe the proportion of HCFs, which receive different services, and to report progressive improvements [4]. A “basic” level of WASH services corresponds to the minimum combination of WASH services required to protect patients and staff’s health [5].

Subsequently, worldwide, the JMP reports that one in four HCFs lack functional water supply on premises, one in three lack hand hygiene facilities, and one in three lack adequate infectious waste disposal [6]. Insufficient piped water on the HCF premises limits handwashing, performing of safe surgeries or deliveries, and cleaning, leading to an increase prevalence of health care acquired infections, which are two to twenty times more prevalent in low-and middle-income countries than in developed ones [4,7]. The situation is even worse in resource limited settings with 50%, 63%, >25%, and 70% of HCFs lacking access to piped water, sanitation facilities, hand hygiene facilities, and appropriate waste disposal systems respectively and considering the detrimental effects of seasonal water shortages, non-functional water infrastructure, and fluctuating water quality commonly experienced there [2,6,8]. In fact, an evaluation of representative data from six countries revealed that only 2% of HCFs provided all four of the required service—water, sanitation, hygiene, and waste disposal [4].

According to the Ministry of Public Health, in Cameroon, a low middle income country of sub-Saharan Africa experiencing economic water scarcity, mortality related to poor WASH practices is estimated at 45.2 deaths per 100 000 inhabitants [9]. In addition to that, in the last decade, Cameroon has experienced several waves of cholera epidemics with the northern and costal zones of the country being the main foci. Especially, the Littoral Region includes the most affected urban districts while the Far-North Region includes the most affected rural districts at the national level [10]. In 2014, the Ebola epidemic in West Africa highlighted the deathful consequences of the lack of hand washing facilities as a first line of defense for health care professionals [11]. Moreover, the COVID-19 pandemics shed light on the lack of investments, the absence of infrastructures, education and policies related to WASH as well as revealed insufficient investment in health care safety and has brought WASH services as non-negotiable for HCFs [6,8].

During the 2019 World Health Assembly, the resolution on WASH in HCFs was unanimously adopted by members of state of the WHO [8]. Furthermore, the COVID-19 pandemics constrained a reassessment of existing norms in national health systems, emphasizing the critical role of adequate WASH practices in protecting human health throughout infectious disease epidemics, ensuring continuity of essential services, and outlining the need to implement country level policies that would prioritize this essential aspect of healthcare delivery. The 2020 global progress report on WASH in HCFs highlighted major gaps in provision of basic hygiene, sanitation, and water services, hence pointing to the critical need to strengthen national surveillance by integrating WASH indicators and obtain a reliable and representative baseline of WASH conditions in each country [5]. Better data monitoring will help to identify low coverage facilities as well as low-cost solutions to improve the situation. The global targets suggest that 80% of HCFs should meet basic WASH services requirement before 2025 [6]. This study aims to assess the effects of the COVID-19 pandemics on WASH in HCFs of the Far-North Region of Cameroon. Specifically, to determine the proportion of HCFs: (1) meeting basic WASH services, and (2) which, WASH services improved post-COVID-19 in the Far-North Region of Cameroon.

## Materials and Methods

### Study Area

The Far-North Region is one of the 10 regions of Cameroon, bordering the North Region to the south, Chad to the east, and Nigeria to the West. With a population of about 5 104 209 inhabitants in 2022, it is considered among the most populated regions of the country and as well as the most densely populated. The region is divided into 32 Health Districts (HDs). The different HDs include 303 Health Areas and about 421 HCFs. Because there is no internationally accepted typology for HCFs, the classification is mostly country-dependent [4]. HCFs in Cameroon are either governmental or private and can be grouped into five categories, which from the lowest to the highest include: (1) integrated health centers (IHC, headed by a senior nurse) and (2) sub-divisional hospitals (CMA, with a physician among the staff); (3) district hospitals, first reference hospital, which offer complimentary package of activities compared to IHC and CMA; (4) Regional hospitals, in charge of specialized health care at the level of the Region. The second and the first categories are represented by Central and General hospitals that handle cases needing more specialized equipment and knowhow [10].

### Study Design and Sampling

This was a cross sectional analytical study whose data collection took place from March 2^nd^, 2022 to June 28^th^, 2022 targeting HDs of the Far-North Cameroon. To obtain the minimum sample size (n) of 91 HCFs to be visited, we used the Cochran’s Modified Formula for Finite Populations [12]: 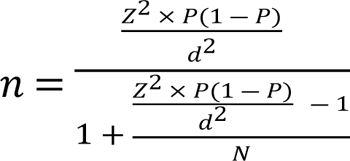 where Z is the approximate value of the 97.5 percentile point of the standard normal distribution =1.96, P is the proportion of adequately functioning WASH services in HCFs=50% (default due to non-availability), d is the precision= 0.1, N=421 (total number of HCFs in all the 32 HDs), and 15% non-response rate.

Due to research constraints, eight HDs: Maroua 1 (21 HCFs), Maroua 2 (17 HCFs), Maroua 3 (17 HCFs), Kousseri (16 HCFs), Makary (08 HCFs), Goulfey (11 HCFs), Mada (13 HCFs) and Fotokol (02 HCFs) were conveniently selected, then proceeded to select all the HCFs in each HD. One respondent per selected HCF was interviewed. From these HDs, HCFs, which managers were absent during data collection, or which were not functional till 12 months pre-COVID-19, or which were located in difficultly accessible geographical areas or areas with high level of insecurity were excluded from the study.

### Data Collection

The study used a validated questionnaire including core indicators that addressed basic WASH services in HCFs [13]. In addition to that, supplementary indicators were used to describe general characteristics of HCFs and respondents and assess the efficacy of WASH services in health care settings. Data collection involved direct observation and discussion with respondents issued from the management team of each selected HCFs. A set of identical questions were asked to obtain a picture of the situation before (12 months pre-COVID-19) and after the COVID-19 pandemics. The survey guide included eight sections: (1) HCFs characteristics (type, category, location, outpatient consultation-only centers); (2) respondents characteristics (function, age, gender, grade); (3) water supply services (type of water source, location of the water source, availability of water); (4) sanitation services (type of latrines, availability of latrines, privacy of latrines, access to people with reduced mobility); (5) hygiene services (availability of hand hygiene facilities at the points of care and near toilets); (6) healthcare waste management services (safe segregation and treatment of wastes); (7) environmental cleaning practices (availability of protocols for cleaning, training of staff with cleaning responsibilities); and (8) other indicators: influence of electricity outages on water availability, water treatment, latrines lightening, sufficient ventilation, gray water drainage, availability of cleaning material, availability of protection equipment for the cleaning staff, and availability of a laundry service [13,14].

### Data Analysis

The data were entered in CSpro (Census and Survey Processing System) version 7.5 and exported to IBM-SPSS version 20 and R version 4.3.0 for statistical analysis. As for descriptive statistics, categorial variables were presented as numbers and percentages whereas numerical variables were presented as means ± standard deviation or median ± interquartile range depending on the distribution. Bivariate analysis was conducted using McNemar test. The significance threshold for p-value was set as <0.05.

Classification of WASH services and its five domains into their corresponding levels per facility was done by calculating the proportion of HCFs providing basic service levels following the JMP guideline [13]. A set of identical questions were asked to describe core indicators frequency and proportions before (12 months pre-COVID-19) and after the COVID-19 pandemics. Individual questions assessing the different indicators for each WASH service i.e., water supply, sanitation, hygiene, waste management, and environmental cleaning were compiled to create the WASH-HCF tool. Following team consensus and literature review based on WASH-FIT and WASH-FAST tools interpretation [13,14], the WASH-HCF tool includes exhaustive indicators of adequate WASH in HCFs. The score for each of the 39 indicators included in the WASH-HCF tool was determined: 0-does not or partially meet the required standards (i.e. target) and 1-fully meets the target. From this, we were able to create an aggregate facility score that can be used to show facilities’ global performance of WASH services and be interpreted as follows: inadequate (total score=0-9), basic (total score=10-19), intermediate (total score=20-29), advanced (total score=30-39). To examine whether the WASH-HCF score differed between the pre-vs. post-COVID-19 period, the test for marginal homogeneity as well as the exact tests of symmetry, including pairwise McNemar test were conducted.

### Ethical Considerations

Ethical clearance N° 3274 CEI-UDo/06/2022/T was obtained from the University of Douala committee and authorizations submitted for approval to the Regional Delegation of Public Health Far-North Region as well as to HCFs directors. Prior to the survey, each HCFs chief received an information notice and a consent form. All Participants provided written informed consent. Because this study can reveal sensitive information of HCFs, all the information that could help identifying HCFs were removed.

## Results

### General characteristics of participants

A total of 97 (23. 04%) HCFs among the 421 that are found in the Far-North region were surveyed for this study (**Table 1**). They were located in eight (25%) of the 32 HDs, were mainly from public management (80.4%), and corresponded in majority to the category of integrated health centers (79.4%). More than half (58.8%) of the HCFs were located in rural vs. urban areas and a similar proportion (55.7%) had a WASH focal point among the staff. The respondents, who were generally the chief of the HCF (92.8%) were mainly males (83.5%) and aged on average 41.18 ± 7.13 years old. The respondents’ professional grades were either nurse (51.5%), assistant nurse (39.2%), or physicians (4.1%) and approximately two-thirds of the respondents had a professional experience of 10 years or less. Even though one-third of respondents were also WASH focal points, only 59% of them reported receiving a WASH-related training.

**Table 1.** General characteristics of HCFs and socio-demographic characteristics of respondents (n=97)

| HCFs characteristics | Numbers(%) | Respondents' characteristics | Numbers(%) |
| --- | --- | --- | --- |
| **District** |  | **Gender** |  |
| Kousseri | 14(14.4) | Male | 81(83.5) |
| Mada | 13(13.4) | Female | 16(16.5) |
| Makary | 8(8.2) | **Age (years)** |  |
| Fotokol | 2(2.1) | ]25-35] | 19(19.6) |
| Goulfey | 11(11.3) | ]35-45] | 47(48.5) |
| Maroua 1 | 18(18.6) | ]45-55] | 27(27.8) |
| Maroua 2 | 16(16.5) | ]55-65] | 4(4.1) |
| Maroua 3 | 15(15.5) | **Position in the facility** |  |
| **Type** |  | Chief | 90(92.8) |
| Public | 78(80.4) | General Supervisor | 7(7.2) |
| Private | 12(12.4) | **Grade** |  |
| Private confessional | 7(7.2) | Medical Doctor | 4(4.1) |
| **Category** |  | Nurse | 50(51.5) |
| Integrated health centers | 77(79.4) | Assistant nurse | 38(39.2) |
| Sub-divisional hospitals | 14(14.4) | Lab assistant | 2(2.1) |
| District hospitals | 4(4.1) | Pharmacy agent | 3(3.1) |
| Regional hospital | 2(2.1) | **Years of experience** |  |
| **Location** |  | <10 | 67(69.1) |
| Urban area | 40(41.2) | [10-20[ | 25(25.8) |
| Rural Area | 57(58.8) | [20-30[ | 4(4.1) |
| **Outpatient-only consultations** | 45(46.4) | ≥30 | 1(1.0) |
| **Absence of usable toilets**<br>(available, functional, private) | 13(13.4) | **The respondent is the**<br>**WASH focal point** | 33(34.0) |
| **Presence of a WASH focal**<br>**point** | 54(55.7) | **The respondent received**<br>**a WASH training** | 57(58.8) |
| **Presence of a funding agency** | 29(29.9) |  |  |

### Availability of WASH services in HCFs

Although 95.9% of HCFs had improved water within 500 m of walking distance, only 77.3% of these water sources were located on premises i.e., within the building or the facility grounds. As well, three-quarter of the HCFs in the study area provided basic water services (**Table 2**). Thirteen HCFs (13.4%) lacked toilets that were available, functional, and private. The median number of usable toilets was 2 (IQR: 2-4). Eighty to ninety percent of the visited HCFs lacked either toilets accessible for people with reduced mobility or for menstrual hygiene management. None of the HCFs met basic sanitation services criteria for healthcare, i.e., by definition, the proportion of HCFs with improved toilets which are usable, sex-separated, provide for menstrual hygiene management, separate for patients and staff, and accessible for people with limited mobility.

**Table 2.**
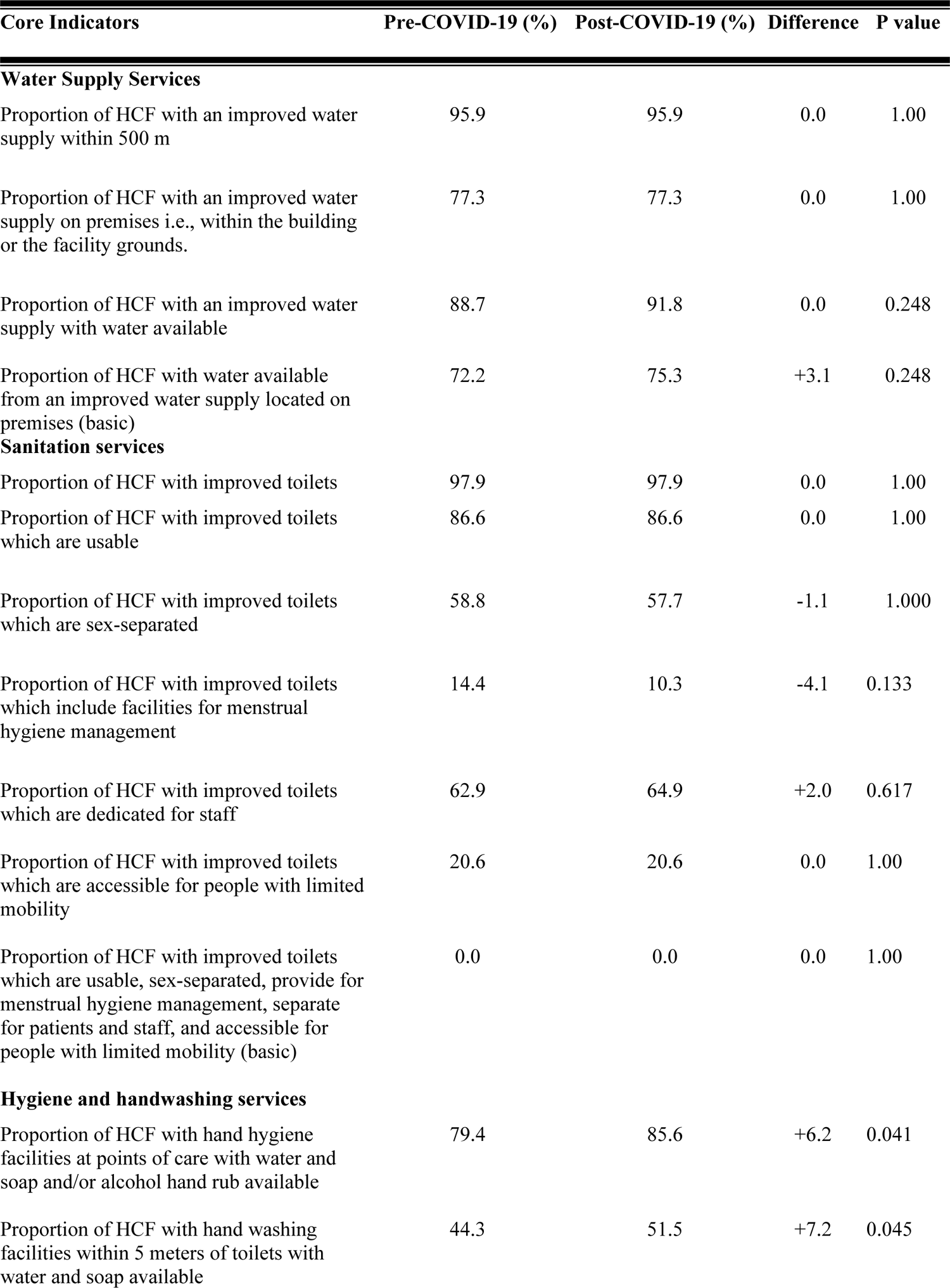
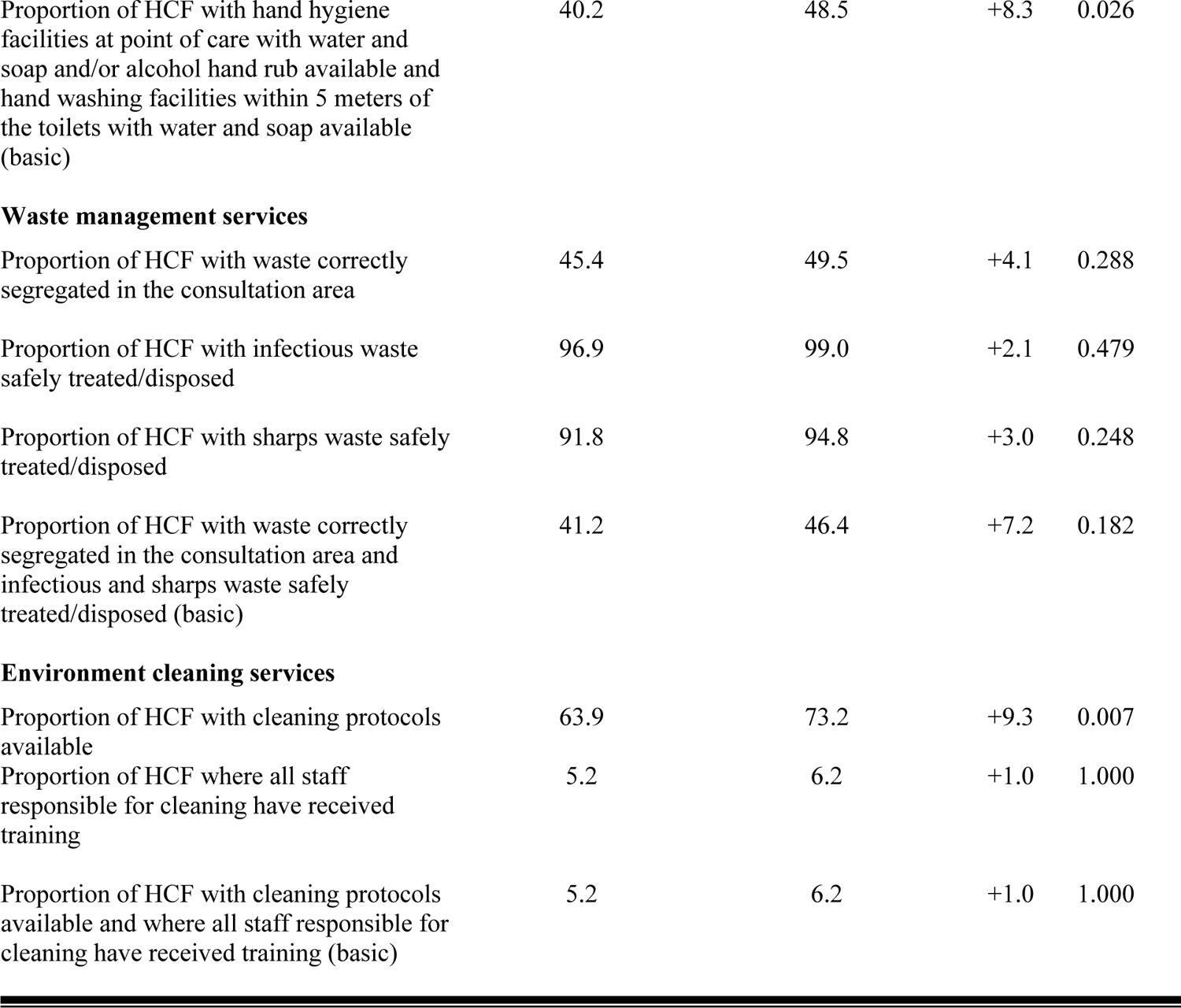
Calculating WASH in HCFs service levels based on responses to the core questions (n=97)

| Core Indicators | Pre-COVID-19 (%) | Post-COVID-19 (%) | Difference | P value |
| --- | --- | --- | --- | --- |
| **Water Supply Services** |  |  |  |  |
| Proportion of HCF with an improved water supply within 500 m | 95.9 | 95.9 | 0.0 | 1.00 |
| Proportion of HCF with an improved water supply on premises i.e., within the building or the facility grounds. | 77.3 | 77.3 | 0.0 | 1.00 |
| Proportion of HCF with an improved water supply with water available | 88.7 | 91.8 | 0.0 | 0.248 |
| Proportion of HCF with water available from an improved water supply located on premises (basic) | 72.2 | 75.3 | +3.1 | 0.248 |
| **Sanitation services** |  |  |  |  |
| Proportion of HCF with improved toilets | 97.9 | 97.9 | 0.0 | 1.00 |
| Proportion of HCF with improved toilets which are usable | 86.6 | 86.6 | 0.0 | 1.00 |
| Proportion of HCF with improved toilets which are sex-separated | 58.8 | 57.7 | -1.1 | 1.000 |
| Proportion of HCF with improved toilets which include facilities for menstrual hygiene management | 14.4 | 10.3 | -4.1 | 0.133 |
| Proportion of HCF with improved toilets which are dedicated for staff | 62.9 | 64.9 | +2.0 | 0.617 |
| Proportion of HCF with improved toilets which are accessible for people with limited mobility | 20.6 | 20.6 | 0.0 | 1.00 |
| Proportion of HCF with improved toilets which are usable, sex-separated, provide for menstrual hygiene management, separate for patients and staff, and accessible for people with limited mobility (basic) | 0.0 | 0.0 | 0.0 | 1.00 |
| **Hygiene and handwashing services** |  |  |  |  |
| Proportion of HCF with hand hygiene facilities at points of care with water and soap and/or alcohol hand rub available | 79.4 | 85.6 | +6.2 | 0.041 |
| Proportion of HCF with hand washing facilities within 5 meters of toilets with water and soap available | 44.3 | 51.5 | +7.2 | 0.045 |
| Proportion of HCF with hand hygiene facilities at point of care with water and soap and/or alcohol hand rub available and hand washing facilities within 5 meters of the toilets with water and soap available (basic) | 40.2 | 48.5 | +8.3 | 0.026 |
| **Waste management services** |  |  |  |  |
| Proportion of HCF with waste correctly segregated in the consultation area | 45.4 | 49.5 | +4.1 | 0.288 |
| Proportion of HCF with infectious waste safely treated/disposed | 96.9 | 99.0 | +2.1 | 0.479 |
| Proportion of HCF with sharps waste safely treated/disposed | 91.8 | 94.8 | +3.0 | 0.248 |
| Proportion of HCF with waste correctly segregated in the consultation area and infectious and sharps waste safely treated/disposed (basic) | 41.2 | 46.4 | +7.2 | 0.182 |
| **Environment cleaning services** |  |  |  |  |
| Proportion of HCF with cleaning protocols available | 63.9 | 73.2 | +9.3 | 0.007 |
| Proportion of HCF where all staff responsible for cleaning have received training | 5.2 | 6.2 | +1.0 | 1.000 |
| Proportion of HCF with cleaning protocols available and where all staff responsible for cleaning have received training (basic) | 5.2 | 6.2 | +1.0 | 1.000 |

Approximately 85.6% of HCFs had functional hand hygiene facilities at points of care compared to 51.1% within 5 m of the toilets. Less than half (48.5%) of HCFs met basic levels for hygiene and handwashing services. A similar proportion of HCFs (46.4%) had waste correctly segregated in the consultation area and infectious and sharps waste safely treated/disposed. As for environmental cleaning services, even though 73.2% of HCFs had cleaning protocols that included step-by-step techniques for different cleaning tasks as well as a cleaning schedule specifying both the responsibilities and the frequency of those tasks, only 6% of the staff responsible for cleaning had received any sort of training (Table 2).

### WASH service levels based on responses to core questions pre-and post-COVID-19

There was no significant change in the proportion of HCFs with improved water supply either located on premises or within 500 m, 12 months pre-COVID-19 vs. the post-COVID-19 period. However, even though not statistically significant (p=0.248), there was an increase (∼+3%) in the percentage of HCFs with basic water access (Table 2). As for sanitation services, the proportion of improved toilets with special commodities (sex-separated, including facilities for menstrual hygiene management) decreased, but the difference in pre- and post-COVID-19 proportions was not significant.

A significant increase was noted in the proportion of HCFs as of optimal handwashing practices related services. Especially, an increase of 8% was noted in a basic hygiene service prior to the COVID-19 pandemics compare to the post-COVID-19 period (P=0.0026). The proportion of HCFs with correctly segregated waste passed from 45.36% to 49.48% pre- and post-COVID-19 respectively. Also, there was an increase in the proportion of HCFs with basic waste management services. But, the difference was not statistically significant. There was a significant increase (p=0.007) in the proportion of HCFs with cleaning protocols available. However, no significant difference was found in the percentage of HCFs with basic environment cleaning services (Table 3).

**Table 3.**
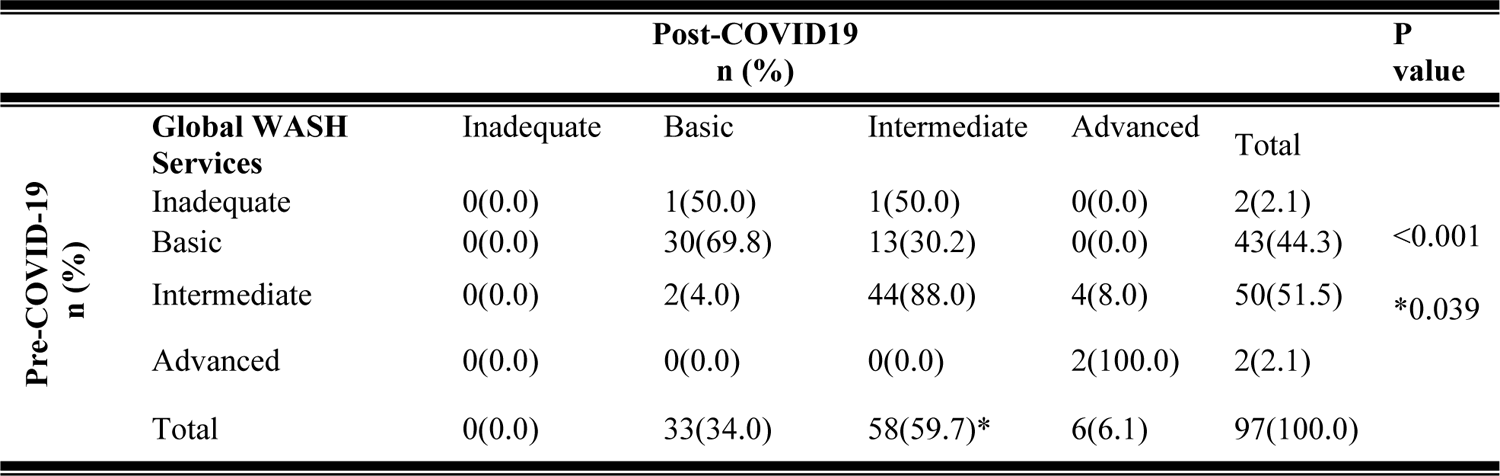
Assessment of the differences in global WASH services pre- and post-COVID-19 using the WASH-HCF tool scores (n=97).

When all five domains were considered together, none of the investigated HCFs fulfilled all criteria to meet basic services. When excluding sanitation and environment cleaning services, there was a 3% not significant increase pre-vs. post-COVID-19 periods. However, when focusing on water and hygiene and handwashing domains, there was an 8% increase between the proportion of HCFs meeting basic criteria for both services pre- and post-COVID-19, with this difference being significative (p=0.045). On the other hand, the proportion of HCFs, which did not meet basic levels for any of the five WASH services decreased from 11.3% to 7.2% and the difference was not significant. The evolution of a HCF before compared to after COVID-19 represented the independent variable and was measured as the proportion of HCFs, which situation evolved in any of the WASH services basic levels. This proportion for water, sanitation, hygiene, waste management, and environmental cleaning was respectively 3.1%, 0.0%, 9.3%, 7.2%, 1% (Table 3). Overall, 13.4% of HCFs evolved pre- to post-COVID-19 period.

### Use of the WASH-HCF Tool

When assessing the differences in global WASH services using the WASH-HCF score for each HCF, the minimum score passed from 7 pre-COVID-19 to 10 post-COVID-19 and the mean score passed from 20.31±5.32 to 21.08±4.82. There was a significant change in the distribution of HCFs according to their global WASH service levels pre-vs. post-COVID-19 (p<0.001). In addition to that, as shown in **Table 3**, all the two HCFs that showed inadequate global WASH services pre-COVID-19 evolved to basic (50%) and intermediate (50%) levels. Approximately 30% of HCFs, which showed basic global WASH services pre-COVID-19 evolved to the intermediate level post-COVID-19 ((p=0.039). The majority of HCFs (88%) from the intermediate level as well as those from the advance level (100%) did not change their category pre vs. post-COVID-19. Only two HCFs, initially in the intermediate category, regressed to the basic level after the COVID-19 pandemics (Table 3). The evolution of the HCFs was also measured based on their WASH-HCF scores: there was 27.8% HCFs, which scores increased from pre- to post-COVID-19 period.

## Discussion

An increased access to WASH services in HCFs is needed in poor settings not only to ensure staff and patient safety, but also to improve quality of care and strengthen global health security [15]. In this study, the response to the COVID-19 pandemics partially improved WASH services-related infrastructures in HCFs of the Far-North Region of Cameroon. Especially, even though there was no significant difference in basic water and sanitation services access, there was a significant increase in HCFs meeting basic hygiene services as well as in the proportion of HCFs with cleaning protocols available.

A safe, adequate, and consistent access to water in HCFs is vital for reducing the transmission and morbidity of infectious diseases [16]. In this study, 95.9% of HCFs had improved water within 500 m of walking distance, 77.3% of these water sources were located on premises. This is similar to the results of a study, which found that 88% of health facilities across 18 countries in sub-Saharan Africa had improved water sources on premises between 2013-2018 [17]. However, it is very important to understand that the presence of water sources does not always translate into regular access to patients and visitors [18]. Research on household water insecurity has demonstrated that beyond objective indicators such as presence/absence of improved water source, distance to the water source, a robust measurement needed to encompass daily water use and intake at the household and at the individual levels [19]. When focusing on basic water access, the study reveals a percentage of 75.3%, lower to what found in Ethiopia in 2022 (88%) [16] and higher than the sub-Saharan Africa 2016 estimates of 74% [17] as well as the 52.4% estimates found in a study in 14 low and middle income countries [20].

The presence of HCFs meeting basic handwashing services represents one of the most critical preventive measures against the transmission of infectious diseases [16]. Healthcare associated infections are 2-20 times higher in low- and middle-income countries compared to developed countries and affect between 2-15% of hospital patients [7]. The WHO recommends functional hand hygiene facilities at all critical points of the HCF and due to the COVID-19 pandemics, hand hygiene awareness has gained great momentum worldwide [16,21]. Approximately 48.5% of HCFs in this study had functional handwashing stations both near latrines and at point of care, which was lower than the global target of 80% [6], but twice higher than the percentage found in Ethiopia in 2022 [16]. However, despite this low proportion, hand washing services indicators were significantly improved by the COVID-19 pandemics as well as the proportion of HCFs with cleaning protocols available. This could be because during the COVID-19 pandemics, there was a mandatory increase in handwashing stations installation as well as the availability of soap and hydroalcoholic solutions. In this study, only 6% of the staff responsible for cleaning had received any sort of training, highlighting the need for an emphasis to be put on training and education. In fact, as a study in Rwanda reported frequent misuse and/or theft of handwashing facilities [16] and adequate WASH in HCF could set examples for proper hand hygiene behavior to be carried into households and communities [20].

Some limitations of this study include the retrospective collection of data that could induce memory bias from the respondents and the small sample size that could preclude the generalizability of the results. Moreover, lack of laboratory analysis of water supply samples may have overestimated the number of HCFs with basic access to water. This study used a tool with 39 validated indicators that could facilitate comprehensive assessment of WASH services in HCFs and hence, through the obtained score, allow the identification of health care centers that require priority in action by the government. More research could be conducted to adapt the indicators by ward or by medical specialty withing each HCF and also to extend the use of the WASH-HCF tool to other regions of Cameroon and even of sub-Saharan Africa.

## Conclusions

The COVID-19 pandemics was a missed opportunity in the study area to provide sufficient water for patients and health workers, build disability-friendly sanitation facilities, and more handwashing stations at point of care and near latrines. There should be a continuing encouragement of governments and funding agencies in planning and budgeting WASH in healthcare-related research and issues, and enable the maintenance of existing WASH infrastructures in healthcare settings. Especially, acknowledging the low proportion of WASH trained staff, the implementation of a WASH committee in HCF including both health care personnel and community members, may increase national recognition of WASH in HCF and its incorporation into policies and have further-reaching effects of WASH-behavior change at the population level. Despite the global chaos the COVID-19 represents, it taught us basic hygiene rules, low risk management of WASH services, further strengthening healthcare acquired infections prevention.

## Data Availability

The data are held in a public repository: http://dx.doi.org/10.13140/RG.2.2.12876.18562

## Acknowledgements

The authors are thankful to the health care facilities authorities for participating in the study.

